# Federated Learning Performance Depends on Site Variation in Global HIV Data Consortia

**DOI:** 10.64898/2026.03.25.26349286

**Authors:** Nicholas J. Jackson, Chao Yan, Yanink Caro-Vega, Fabio Paredes, Ronaldo Ismerio Moreira, Stanley Cadet, Diana Varela, Carina Cesar, Stephany N Duda, Bryan E. Shepherd, Bradley A. Malin

**Author notes:** **Contact author and address:** Nicholas J Jackson, Address: 2525 West End Ave, Suite 1475, Nashville, TN, 37203-1536.

## Abstract

Digital health technologies, including machine learning (ML), are transforming infectious disease management, however ML models for HIV care have been limited by data sharing restrictions that prevent multi-site collaboration. Federated Learning (FL) offers a privacy-preserving solution, enabling cross-site model training without sharing patient-level data. We evaluated FL for developing clinical prediction models using data from 22,234 people living with HIV (PLWH) across six sites in five countries within the Caribbean, Central, and South America network for HIV epidemiology (CCASAnet). Across four prediction tasks — 1-year mortality, 3-year mortality, tuberculosis incidence, and AIDS-defining cancer incidence — FL algorithms achieved near-centralized performance while substantially outperforming site-specific models. Performance gains varied across sites, driven by both site size and between-site heterogeneity. Local fine-tuning often improved FL performance, though benefits were task dependent. These findings support FL as a scalable, privacy-preserving infrastructure for multi-site ML in international HIV research.

## Introduction

The rapid advancement of digital health technologies is reshaping the prevention, surveillance, and clinical management of infectious diseases, with artificial intelligence and machine learning (ML) emerging as powerful tools for population-level disease surveillance^1^ or aiding in clinical decision-making^2^. For people living with HIV (PLWH), of whom there are an estimated 40 million worldwide, ML models offer a range of opportunities. For example, ML models applied to HIV surveillance data can track epidemic trajectories and inform resource allocation at the population level^3^, while models embedded in routine clinical practice can forecast key events in the HIV care continuum — including predicting mortality,^4^ incident tuberculosis,^5^ AIDS-defining cancers^6^, and identifying PLWH at risk for treatment failure^7^ —allowing medical professionals to more effectively allocate scarce healthcare resources.^8^

However, realizing this potential requires ML models to be trained on large, diverse datasets across varied geographies, health systems, and patient populations — datasets that are difficult to assemble in practice. As such, present research has been limited to single-site datasets, often with small sample sizes.^9^ This limitation is critical as ML models require large amounts of training data, and small datasets may be insufficient to train reliable and generalizable models. This limitation potentially leaves many PLWH, particularly those in low-resource settings, without access to the benefits of ML technology. Historically, public health institutions and research programs have overcome similar barriers by collaborating and pooling data from multiple cohorts to create large, diverse datasets.^10,11^ While some researchers have leveraged these datasets for the development of ML models,^12^ the creation and maintenance of these large collaborative datasets comes with significant challenges: they are costly and time intensive to establish, and when collaborations span national borders, data sharing is frequently constrained by privacy or data-sharing regulations.^13,14^

Federated Learning (FL) offers a promising solution to these challenges by enabling cross-site ML model training without sharing raw, patient-level data. Instead, each site retains its data locally and only shares model updates (e.g., model parameters) that are aggregated to build a shared (global) model. This privacy-preserving architecture directly addresses the data governance and regulatory barriers that have historically limited multi-site ML development in international health research.^15^ FL has shown impressive performance in many different medical ML tasks, often achieving performance comparable to models trained using centralized datasets.^16–18^ However, a potential limitation of FL models is that they are sensitive to heterogeneity between sites; differences in the patient populations across sites can disrupt the learning process of FL models, potentially rendering them ineffective. In the HIV domain, such heterogeneity arises from differences in healthcare access,^19^ the prevalence of HIV and opportunistic infections,^20^ local clinical practices,^21^ and data standardization practices.^22^

Despite the clear promise of FL as a solution for privacy-preserving multi-site ML, to the best of our knowledge, it has not yet been evaluated in the context of HIV. Thus, we systematically evaluated the efficacy of FL for developing clinical prediction models using a cohort of over 26,000 PLWH derived from the Caribbean, Central, and South America network for HIV epidemiology (CCASAnet),^10^ a large and diverse international HIV consortium spanning multiple countries across Latin America and the Caribbean. We investigated four common FL algorithms and compared their performance to centralized ML training algorithms (i.e., unrestricted data sharing) and site-specific models (i.e., no data sharing). To ensure generalizability, we developed models for four different clinical prediction tasks and evaluated model performance across six clinical sites from five countries. Beyond benchmarking performance, we conducted ablation experiments to characterize the conditions under which FL succeeds or fails, empirically illustrating how between-site heterogeneity and site size shape FL utility, with direct implications for the design and deployment of FL systems in real-world, resource-diverse settings.

## Results

### Overview

We evaluated seven ML training approaches across three data-sharing scenarios—Centralized (unrestricted data sharing), Site-Specific (no data sharing), and Federated Learning (FL; no sharing of patient-level data)—for four clinical prediction tasks: 1-year mortality, 3-year mortality, 1-year tuberculosis incidence, and 1-year AIDS-defining cancer incidence. FL was implemented using two algorithms (FedAvg and FedProx) and their locally fine-tuned counterparts (FedAvg-FT and FedProx-FT), with Centralized and Centralized-FT models serving as upper-bound benchmarks and Site-Specific models as a lower-bound baseline. All models were fully connected neural networks evaluated on data from 22,234 PLWH across six CCASAnet sites in five countries; demographic and clinical characteristics of the cohort are summarized in **Table 1**. Performance was measured primarily by area under the receiver operating characteristic curve (AUC). Results for other performance metrics (F1, sensitivity, specificity) are available in Supplementary Figures 1-3. We find that FL approaches centralized performance across all four tasks, though performance gains varied substantially across sites, a pattern we attribute to both site size and between-site heterogeneity.

**Table 1.** Demographic and basic clinical characteristics of the six CCASAnet sites at ART initiation with a least 1-year follow-up. PLWH: People with HIV. Honduras-1, Honduras-2, and Haiti were omitted from the cancer prediction task as they did not collect data on cancer outcomes (denoted “-“).

|  | Overall | Brazil | Chile | Honduras-1 | Honduras-2 | Haiti | Mexico |
| --- | --- | --- | --- | --- | --- | --- | --- |
| **Number of PLWH** | 22,234 | 4,390 | 2,299 | 382 | 283 | 13,456 | 1,424 |
| **Sex**<br>**n (%)** |  |  |  |  |  |  |  |
| **Female** | 9,797<br>(44.1) | 1,233<br>(28.1) | 251<br>(10.9) | 169<br>(44.2) | 80<br>(28.3) | 7,947<br>(59.1) | 117<br>(8.2) |
| **Male** | 12,411<br>(55.8) | 3,157<br>(71.9) | 2,022<br>(88.0) | 213<br>(55.8) | 203<br>(71.7) | 5,509<br>(40.9) | 1,307<br>(91.8) |
| **Unknown** | 26<br>(0.1) |  | 26<br>(1.1) |  |  |  |  |
| **Age**<br>**mean (SD)** | 37.2<br>(10.9) | 36.1<br>(10.6) | 34.8<br>(10.1) | 34.7<br>(9.6) | 39.6<br>(10.6) | 38.3<br>(11.1) | 35.1<br>(10.1) |
| **BMI**<br>**mean (SD)** | 20.5<br>(2.3) | 20.8<br>(1.5) | 20.0<br>(0.7) | 21.7<br>(2.6) | 21.1<br>(1.0) | 20.4<br>(2.5) | 21.9<br>(3.4) |
| **CD4 cell count**<br>**mean (SD)** | 266.1<br>(203.0) | 307.8<br>(229.1) | 256.2<br>(174.1) | 176.5<br>(149.5) | 175.4<br>(134.2) | 261.3<br>(197.7) | 241.7<br>(209.3) |
| **Log<sub>10</sub> HIV Viral**<br>**Load**<br>**mean (SD)** | 10.5<br>(1.8) | 10.4<br>(2.0) | 10.7<br>(1.9) | 10.4<br>(2.6) | 10.4<br>(1.4) | 10.5<br>(1.7) | 11.2<br>(1.9) |
| **Death (1-year)**<br>**n (%)** |  |  |  |  |  |  |  |
| **No** | 21,328<br>(95.9) | 4,237<br>(96.5) | 2,235<br>(97.2) | 371<br>(97.1) | 270<br>(95.4) | 12,823<br>(95.3) | 1,392<br>(97.8) |
| **Yes** | 906<br>(4.1) | 153<br>(3.5) | 64<br>(2.8) | 11<br>(2.9) | 13<br>(4.6) | 633<br>(4.7) | 32<br>(2.2) |
| **Death (3-year)**<br>**n (%)** |  |  |  |  |  |  |  |
| **Excluded due to**<br>**< 3-year follow-up** | 4,521<br>(20.3) | 107<br>(2.4) | 624<br>(27.1) | 33<br>(8.6) | 34<br>(12.0) | 3,626<br>(26.9) | 97<br>(6.8) |
| **No** | 16,251<br>(73.1) | 4,039<br>(92.0) | 1,569<br>(68.2) | 326<br>(85.3) | 229<br>(80.9) | 8,812<br>(65.5) | 1,276<br>(89.6) |
| **Yes** | 1,462<br>(6.6) | 244<br>(5.6) | 106<br>(4.6) | 23<br>(6.0) | 20<br>(7.1) | 1,018<br>(7.6) | 51<br>(3.6) |
| **TB**<br>**n (%)** |  |  |  |  |  |  |  |
| **Excluded due to**<br>**prior TB** | 2,093<br>(9.4) | 599<br>(13.6) | 44<br>(1.9) | 18<br>(4.7) | 12<br>(4.2) | 1,328<br>(9.9) | 92<br>(6.5) |
| **No** | 19,479<br>(87.6) | 3,627<br>(82.6) | 2,231<br>(97.0) | 351<br>(91.9) | 260<br>(91.9) | 11,709<br>(87.0) | 1,301<br>(91.4) |
| **Yes** | 662<br>(3.0) | 164<br>(3.7) | 24<br>(1.0) | 13<br>(3.4) | 11<br>(3.9) | 419<br>(3.1) | 31<br>(2.2) |
| **Cancer**<br>**n (%)** |  |  |  |  |  |  |  |
| **Excluded due to**<br>**prior Cancer** | 204<br>(0.9) | 92<br>(2.1) | 43<br>(1.9) | - | - | - | 66<br>(4.6) |
| **No** | 21,820<br>(98.1) | 4,207<br>(95.8) | 2,190<br>(95.3) | - | - | - | 1,308<br>(91.9) |
| **Yes** | 210<br>(0.9) | 91<br>(2.1) | 66<br>(2.9) | - | - | - | 50<br>(3.5) |

### FL achieves near-centralized performance without patient-level data sharing

Across all four prediction tasks, FL algorithms performed nearly as well as Centralized models while substantially outperforming Site-Specific models (**Table 2**). The best-performing FL algorithm, FedProx-FT, achieved AUCs of 0.758, 0.744, 0.844, and 0.784 for 1-year mortality, 3-year mortality, tuberculosis, and cancer prediction respectively, compared to 0.762, 0.743, 0.848, and 0.779 for the best-performing Centralized approach and 0.747, 0.736, 0.831, and 0.765 for Site-Specific models. In general, Centralized model training achieved the highest AUC, outperforming all other methods when predicting 1-year mortality and 1-year cancer incidence. Notably, fine-tuning algorithms (Centralized-FT, FedAvg-FT, and FedProx-FT) often performed comparably to or better than their counterparts without fine-tuning (Centralized, FedAvg, and FedProx, respectively). Specifically, Centralized-FT outperformed Centralized algorithms in 3-year mortality. Fine-tuning often improved federated algorithms as well, with FedAvg-FT and FedProx-FT outperforming their respective counterparts for both mortality prediction tasks and the tuberculosis prediction task (**Table 2**). As expected, Site-Specific models were the least effective across all four tasks.

**Table 2.** Overall performance (measured using AUC) of different training approaches averaged over 250 runs (± Standard Error). Site-Specific is the worst-case baseline where each site only trains on its own data. The best performing federated methods were bolded. The best-performing baselines (i.e., non-FL approaches where all data are available for training) are underlined. The best-performing algorithms for each task are marked with asterisks (*).

| Model | 1-Year Mortality<br>AUC ± SE | 3-Year Mortality<br>AUC ± SE | 1-Year Cancer<br>AUC ± SE | 1-Year Tuberculosis<br>AUC ± SE |
| --- | --- | --- | --- | --- |
| Site-Specific | 0.747 ± 0.002 | 0.736 ± 0.001 | 0.831 ± 0.002 | 0.765 ± 0.002 |
| FedAvg | 0.757 ± 0.001 | 0.736 ± 0.001 | **0.846 ± 0.002** | 0.774 ± 0.001 |
| FedProx | 0.754 ± 0.001 | 0.737 ± 0.001 | **0.846 ± 0.002** | 0.776 ± 0.001 |
| FedAvg-FT | **0.758 ± 0.001** | 0.742 ± 0.001 | 0.841 ± 0.002 | 0.781 ± 0.001 |
| FedProx-FT | **0.758 ± 0.001** | **0.744 ± 0.001** | 0.844 ± 0.001 | **0.784\* ± 0.001** |
| Centralized | <u>0.762* ± 0.001</u> | 0.743 ± 0.001 | <u>0.848* ± 0.002</u> | <u>0.779 ± 0.001</u> |
| Centralized-FT | 0.759 ± 0.001 | <u>0.747* ± 0.001</u> | 0.844 ± 0.002 | 0.777 ± 0.001 |

### Site size partially explains variation in FL performance

Not all sites benefitted equally from FL. Haiti, the largest site with 13,456 PLWH, saw negligible performance improvements from FL relative to Site-Specific models across all four tasks, while smaller sites such as Mexico (1,424 PLWH) and both Honduras sites (382 and 283 PLWH) saw substantially larger gains (**Figure 1, blue vs purple**). One intuitive explanation is site size; smaller sites have less training data and would be expected to benefit more from FL than larger ones. To isolate this effect, we resampled the CCASAnet data to create six sites with homogenous populations, ensuring that sample size was the only meaningful difference between them. Consistent with our intuition, smaller sites generally showed greater performance improvements under this controlled scenario **(Figure 2, A**). However, the magnitude of this improvement often differed from what was observed in our primary analysis **(Figure 2, B**). For instance, despite having similar sizes, Honduras-1 saw much greater performance improvements in the primary analysis than Honduras-2, and the improvements from these sites did not align with those of their counterpart simulated sites. This suggests that the performance gains within a specific site are not entirely driven by the number of training samples. Results from the other three prediction tasks likewise showed such differences and are available in Supplementary Figures 4-6.

**Figure 1.**
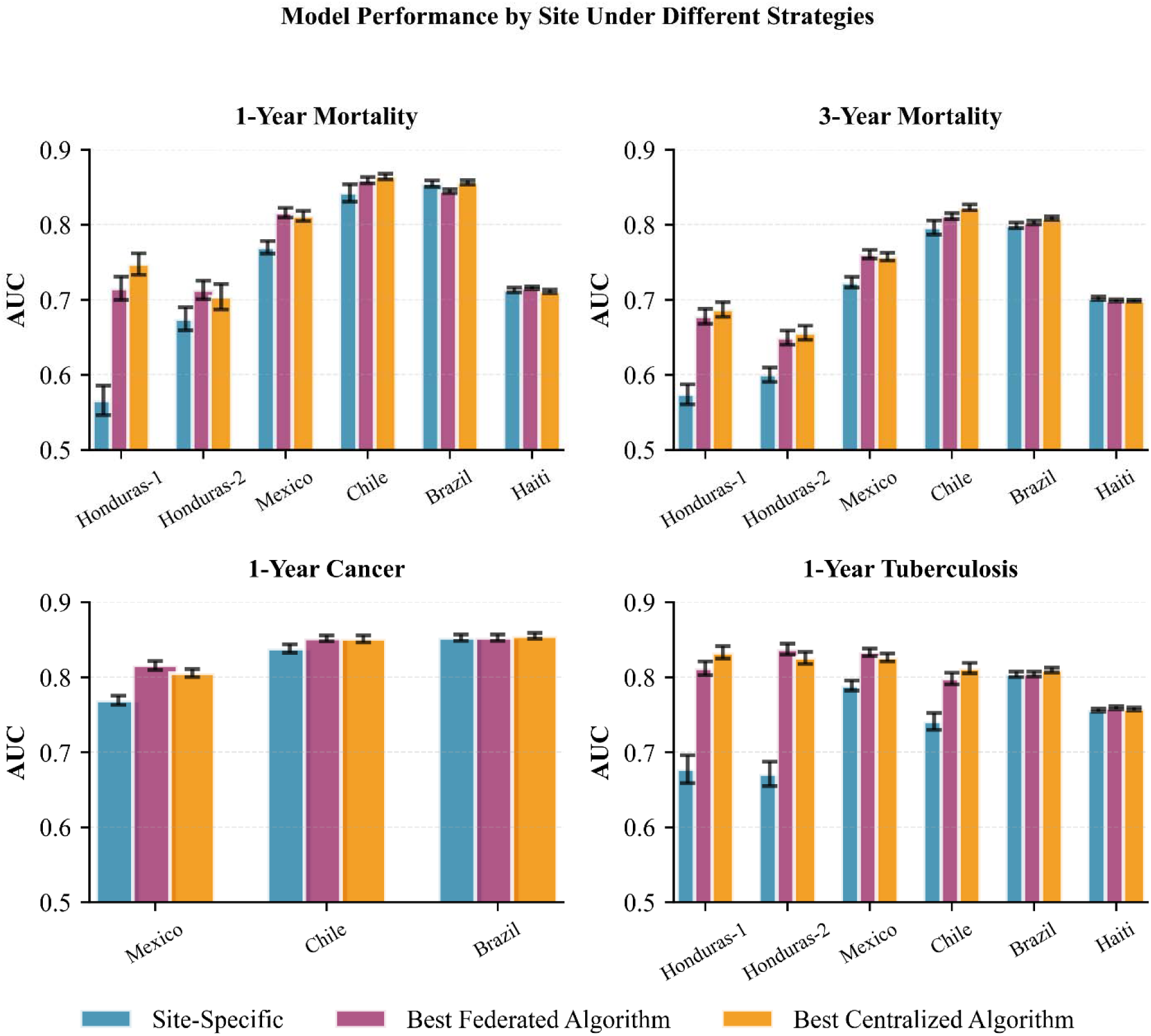
Performance of different ML training approaches on each site for each task. Sites are sorted from left to right in ascending size (i.e., number of PLWH). Centralized algorithms (orange) generally perform the best on each site as there are no data restrictions while Site-Specific (blue) performs the worst on each site as this is the largest restriction on data. Honduras-1, Honduras-2, and Haiti were omitted from the cancer prediction as they did not collect data on cancer outcomes. Error bars represent standard error over 250 repeated experiments.

**Figure 2.**
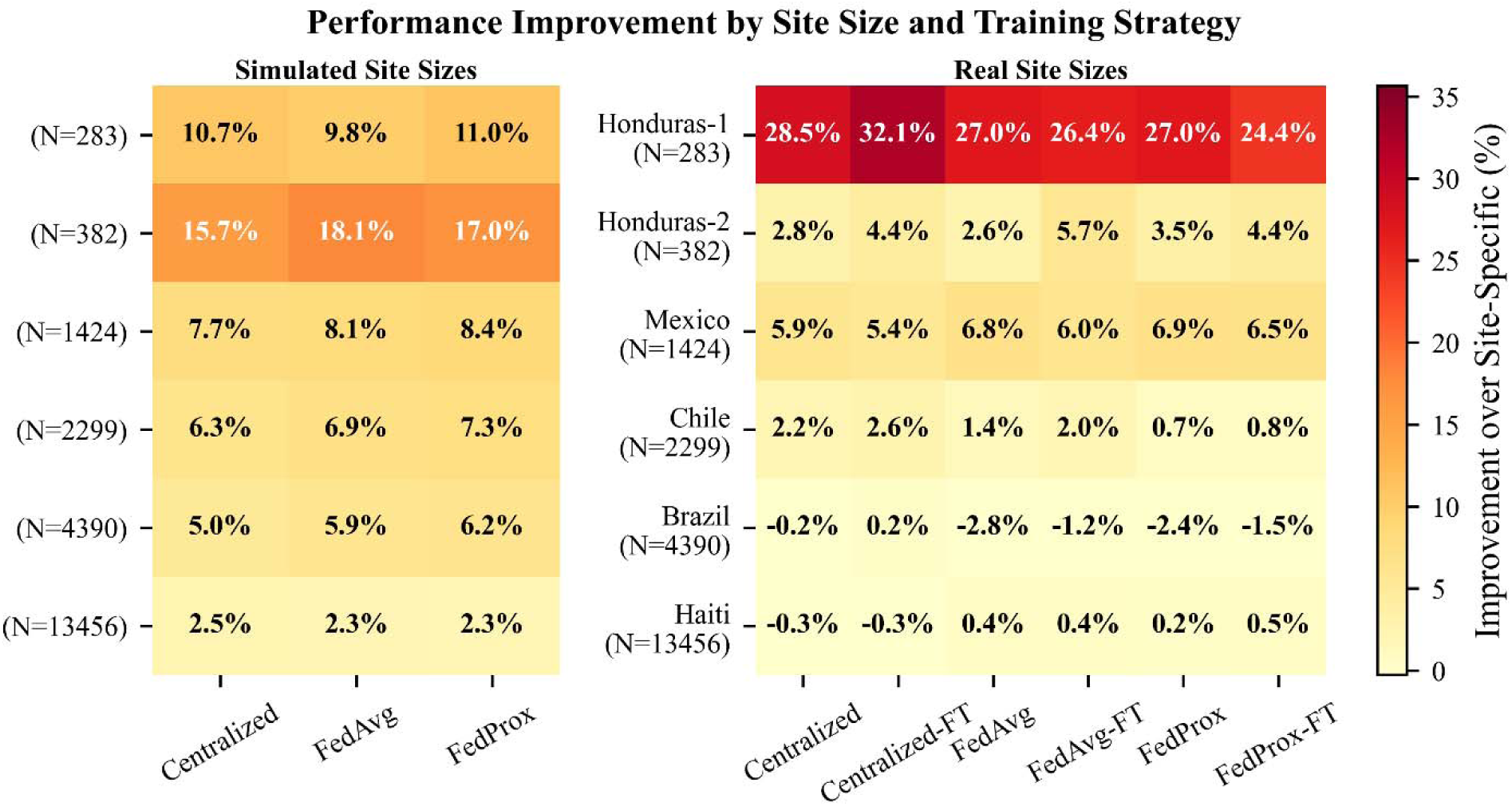
Relative change (%) in AUC compared to Site-Specific models in both the Site Size (left) and Real-World experiments (right) for the 1-year mortality prediction task. Darker Cells indicate the greater increase in AUC. Smaller sites generally had greater performance improvements than larger sites, but these trends did not reflect those observed in the Real-World experiment. As expected, we observed that fine-tuning offered no performance benefits for IID sites (left). As a result, we only present results from the Centralized, FedAvg, and FedProx model training approaches for these experiments.

### Between-site heterogeneity independently drives FL performance

To directly assess the impact of between-site heterogeneity on FL performance, we applied a latent variable clustering algorithm to data from Brazil, generating simulated sites of varying heterogeneity controlled by a parameter α, where higher values of α correspond to greater heterogeneity. **Figure 3** details these results for the cancer prediction task, where we observed that Centralized models performed the best (**Figure 3, blue**), followed by FedProx, FedAvg, and Site-Specific models. While, in general, Site-Specific models were the least performant, they often performed better under high heterogeneity (**Figure 3, pink**). In contrast, performance of both FL algorithms decreased under higher heterogeneity (**Figure 3, green, purple**) such that Site-Specific models were preferable under very high heterogeneity.

**Figure 3.**
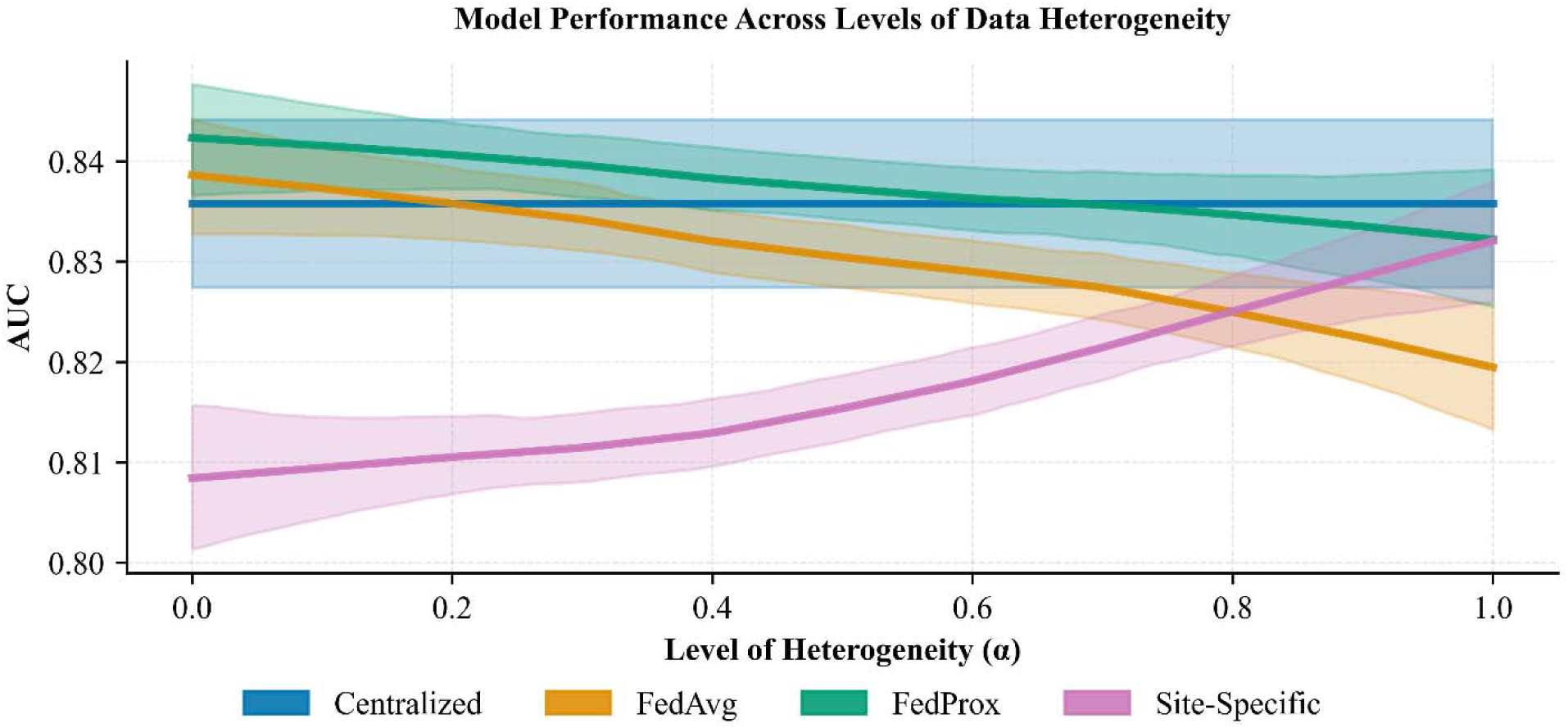
Tuberculosis prediction performance on datasets with simulated heterogeneity between sites. Higher values of α have greater heterogeneity between sites. When α = 0 there is no between-site heterogeneity (i.e., they are IID samples of the population). Centralized (blue) was the most performant methods across the range of α and Site-Specific was generally the least. FL generally performed worse for high heterogeneity (purple, green), however Site-Specific (pink) increased under high heterogeneity, outperforming FedAvg and performing comparably to FedProx. Curves were fit to the raw data using lowess regression; 95% confidence intervals were generated via bootstrap resampling. Results from fine-tuned variants of these models were omitted from the figure as they did not differ substantially from their non-fine-tuned counterparts.

The observation that FedAvg and FedProx worsened under high heterogeneity was consistent for all four prediction tasks; however, the performance of Site-Specific models varied by task, highlighting that each unique prediction task may carry differing levels of heterogeneity. Results for the remaining prediction tasks are available in Supplementary Figures 7-9.

### Local fine-tuning improves FL performance

Across all four prediction tasks, locally fine-tuned FL algorithms consistently matched or outperformed their non-fine-tuned counterparts (Table 2). This benefit was most pronounced for the tuberculosis prediction task, where FedProx-FT outperformed all other approaches including Centralized training. This suggests that fine-tuning allows FL models to recover site-specific patterns that are diluted or obscured during global model aggregation. However, the benefit of fine-tuning was task-dependent, highlighting that the decision to apply fine-tuning should be evaluated on a per-application basis.

## Discussion

Across four clinical prediction tasks spanning mortality, tuberculosis, and AIDS-defining cancer prediction, FL algorithms performed nearly as well as centralized ML models while substantially outperforming site-specific training — and critically, does so without sharing any patient-level data. ML models trained without data-sharing restrictions generally achieved the best performance, as expected, but gaps between such models and FL approaches were small across all four prediction tasks. This finding is significant in the context of international HIV research, where data governance frameworks and privacy regulations frequently prevent the kind of large-scale data sharing that high-performing ML models require. An important caveat is that not all sites benefited equally from FL. Because ML models often require large amounts of data, one might expect that larger sites (i.e., more PLWH) would not benefit much from FL, while smaller sites would benefit the most. While we observed this as a general trend, the relationship between site characteristics and FL performance proved more complex — with between-site heterogeneity emerging as a substantial driver of variation in FL performance across populations.

Building on this, we found that site size generally drives performance gains from FL, consistent with expectations. However, the heterogeneity of the site (i.e., how different its population is from other sites) is also a driving factor. The clearest example of this is Haiti, where Site-Specific models performed nearly as well as Centralized models across all four prediction tasks, in contrast to the more general trend that Site-Specific models were the least performant. This can be attributed to two factors. First, Haiti is the largest site in CCASAnet with over 13,000 PLWH, providing sufficient local data to learn a reliable Site-Specific model without using data from other sites. Additionally, the HIV epidemic in Haiti is distinct from that in other countries with sites participating in CCASAnet, with different data collection practices,^10^ fewer treatment resources,^23^ and an estimated general HIV prevalence several times higher than that in Brazil, Chile, Mexico, and Honduras.^24^ As a result, data from the other CCASAnet sites may be sufficiently distinct that adding these data to the model (via FL or Centralized training) may harm (or at least not improve) model performance in Haiti. This phenomenon, where adding data from different clinical sites reduces model performance, has been observed elsewhere in medical ML and is a topic of emerging research.^25^

Our simulation experiments confirmed that between-site heterogeneity is a primary driver of FL performance (rather than site size alone). While smaller sites benefited the most from FL when heterogeneity was removed, the magnitude of this improvement often differed substantially from our primary analysis on real sites. Furthermore, FedAvg and FedProx both performed worse under high heterogeneity, suggesting that their utility may be limited when sites are highly dissimilar. Collectively, these results indicate that practitioners should carefully assess the similarity of each clinical population before deploying FL.

One simple and practical response to such heterogeneity is local fine-tuning. We found that fine-tuned FL algorithms consistently match or outperformed their non-fine-tuned counterparts across all four tasks. This was most pronounced for tuberculosis prediction, where models trained using Centralized-FT outperformed Centralized models. Similarly, FedAvg and FedProx were both outperformed by their fine-tuned counterparts for the 1-year and 3-year mortality prediction tasks. This suggests that fine-tuned FL methods can, at times, improve performance by focusing on learning site-specific trends that may be missed due to inefficiencies in other FL algorithms. However, this benefit was task-dependent and warrants evaluation for each real-world application.

These findings carry several practical implications for the deployment of FL in international HIV research and beyond. FL is a viable strategy to develop effective predictive ML models, highlighting its potential to address challenges in international data sharing. However, not all sites will benefit equally. The degree of improvement is shaped by both size and between-site heterogeneity such that, larger sites and those with more epidemiologically distinct populations may benefit less from participating in FL than others. For such sites, local fine-tuning offers a practical mitigation strategy and should be considered as a default component of FL pipelines deployed in heterogeneous international collaborations.

This study has several limitations. The FL algorithms used in this study require many communication rounds to converge, which would necessitate dedicated federated infrastructure if deployed in practice. While software exists to facilitate the development of such networks,^26^ one-shot or few-shot federated prediction models may offer a pragmatic path to deployment in low-resource settings.^27^ However, as these methods may be less performant, future work should investigate the efficacy of few-shot federated algorithms to understand how they perform relative to federated ML models such as neural networks and how feasible it is for such analyses to be performed without extensive computational infrastructure (e.g., having statisticians perform the analyses at each site). Beyond infrastructure, data missingness and lack of standardization across sites remain practical barriers to realizing FL’s benefits in practice. As such, future work should develop FL methods that can handle missing data with varied structures and assess their performance on heterogeneous populations. Finally, while our findings are drawn from a large, geographically diverse HIV cohort, their generalizability to other infectious diseases has yet to be established.

In summary, Federated learning offers a compelling path forward for privacy-preserving, multi-site machine learning in international HIV research, achieving near-centralized performance across four clinically meaningful prediction tasks without sharing patient-level data. However, its benefits are not uniform as both between-site heterogeneity and the size of each clinical site governs how much individual sites stand to gain from FL participation. Local fine-tuning offers a practical and effective mitigation strategy for sites with epidemiologically distinct populations. As digital health technologies become increasingly integrated into infectious disease surveillance and clinical care globally, FL represents a scalable and privacy-respecting infrastructure for developing ML models that reflect the diversity of real-world patient populations — particularly in low-resource and international settings where data sharing barriers are most acute.

## Methods

### Data Source and Outcome Definitions

This study used data from CCASAnet, a large international HIV consortium spanning Central America, South America, and the Caribbean, established to support collaborative HIV research across international patient populations. We used data from six HIV care sites across five countries within CCASAnet: Brazil, Chile, Honduras (with two sites), Haiti, and Mexico. We developed models to predict 1-year mortality, 3-year mortality, 1-year incidence of tuberculosis infection, and 1-year incidence of an AIDS-defining cancer (including Kaposi sarcoma, invasive cervical cancer, and non-Hodgkin lymphoma). We included antiretroviral therapy-naïve adults (≥18 years) with HIV who initiated antiretroviral therapy (ART) between 2006-2018 and excluded those lost to follow-up before the end of each task’s prediction window. For the tuberculosis and cancer prediction tasks, PLWH with prior diagnoses of tuberculosis or AIDS-defining cancers were excluded. For the cancer prediction task, we excluded three of the sites (Haiti and both sites from Honduras) as cancer data were not routinely collected at those sites. Covariates were collected within 30 days of ART initiation and included: age, sex, CD4 cell counts, HIV viral load, body mass index (BMI), the presence of AIDS (using AIDS-defining diagnoses), risk factors for HIV acquisition, and years since HIV diagnosis. Multiple imputation (without the use of the predicted outcome) was applied to handle missing data. This study was approved with a waiver of informed consent by each of the CCASAnet sites and the coordinating center at Vanderbilt University Medical Center (IRB #060284).

### Model Training

We compared seven approaches for training ML models: Centralized, Site-Specific, FedAvg, FedProx, as well as variations of Centralized, FedAvg, and FedProx in which copies of each trained model were fine-tuned to each specific site (Centralized-FT, FedAvg-FT, and FedProx-FT, respectively). Each of these approaches to training ML models corresponds to one of three data sharing scenarios.

#### Scenario 1 (Unrestricted Data-Sharing)

In this scenario, patient-level data can be shared freely among sites, and the goal is to develop one model that performs well across all sites.

- Centralized: A single ML model was trained using a standard training procedure, with access to the entire dataset, that iteratively minimizes expected prediction error^28^ (**Figure 4a**). Centralized training is considered the upper limit of performance as no restrictions were imposed on the training of the ML models.
- Centralized with Fine-Tuning (Centralized-FT): Whereas Centralized model development aims to create a single model that is effective for all sites, Centralized-FT aims to develop multiple models, each of which is optimized to a specific site. In this case, the Centralized model was fine-tuned to each of the six sites that contributed data, yielding six ML models. Centralized-FT starts with the model developed via Centralized training before performing fine-tuning (**Figure 4d**). Fine-tuning is performed in the same manner as Centralized model training (i.e., we iteratively minimize prediction error), except it is trained on only one site and with a smaller learning rate and smaller number of iterations (both determined through hyperparameter search).

**Figure 4.**
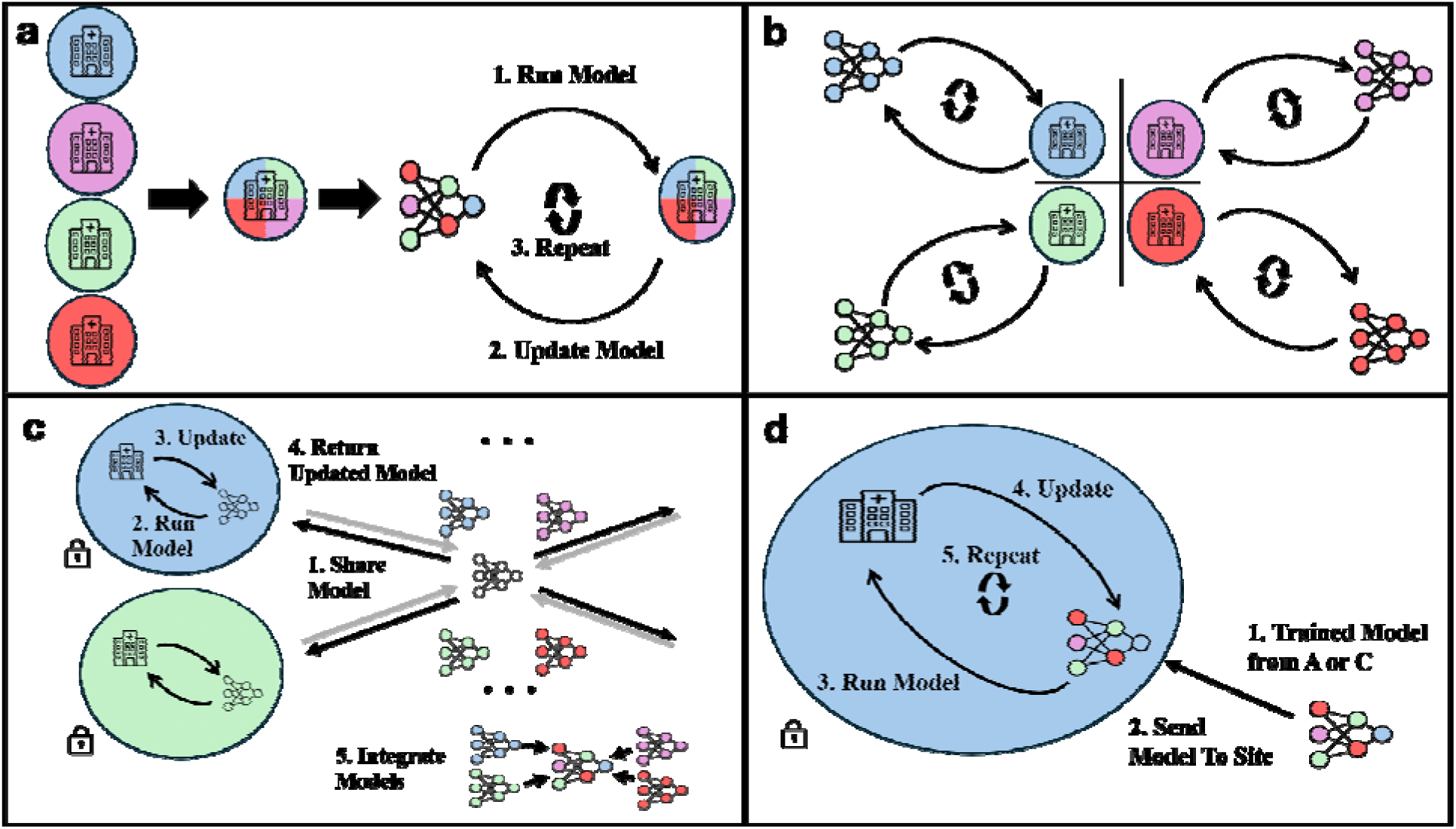
An illustration of the Centralized (**a**), Site-Specific (**b**), standard FL methods such as FedAvg and FedProx (**c**), and both Centralized and FL methods with fine-tuning (**d**). **a**. A Centralized ML model is trained by first aggregating all the data, then training the ML model via iterative updating. **b.** The Site-Specific ML models were developed similarly, except that one model is developed for each site and data are not aggregated as in **a**. **c.** FedAvg and FedProx are both implemented by first creating a global ML model (i.e., by a data coordinating center) that is then sent to each site to be run locally in their secure data environment (black arrow). These models are then updated and sent back to the coordinating center (gray arrow), where they are aggregated into an updated global model. This process is repeated until the model converges. **d.** The model created from either **a.** (Centralized) or **c.** (Federated) is used as a starting point for fine-tuning. After receiving this model, the site iteratively updates the model on their own data until it converges to perform well on their data. This process is conducted for all sites, yielding a unique model for each site.

#### Scenario 2 (No Data-Sharing)

In this scenario, no data are shared across sites. Rather, each site uses only its own data for model training.

Site-Specific: ML models are trained on data from only one site using standard model training procedures^28^ (**Figure 4b**). This case represents the theoretical lower bound on model performance, as one would expect that any additional data available in the other scenarios would improve upon this performance.

#### Scenario 3 (Federated Learning)

In this scenario, patient-level data cannot be freely shared; however, sites participate in a federation where models are trained using FL. To simulate this federation, we partitioned the dataset according to the six sites where data were collected and trained models using the following FL algorithms:

Federated Averaging (*FedAvg*) is one of the most common and simplest FL algorithms (**Figure 4c**).^29^ First, a global model is randomly initialized. Then, at each iteration, a copy of the model is sent to each site where this local model is run, its error is calculated, and the local model is updated based on this error. The global model is then updated as the weighted average (weighted by size of the site) of the local models. This process is repeated until there is no further improvement in the model’s performance.
*FedProx* is similar to FedAvg in that it follows the same iterative procedure described above where it updates local models at each site and then integrates them into a global model. However, FedProx uses a regularization term that minimizes the impact of between-site heterogeneity, thereby limiting noise present in the training process.^30^
Federated Fine-Tuning (*FedAvg-FT* and *FedProx-FT*) aims to use a FL framework to develop the best model for each site. These algorithms are analogous to Centralized-FT, with the only difference being that these algorithms start with the model developed via FedAvg or FedProx instead of the Centralized model (**Figure 4d**).

The ML models in this study were fully connected neural networks and were trained using the cross-entropy loss function. The number of hidden layers in the neural-network models, the width of the layers, the learning rates applied to train the models, and the number of training iterations were tuned using 100 iterations of a Bayesian hyperparameter search.^31^

## Experiments

To evaluate FL performance under real-world conditions, we partitioned the dataset according to the sites where data were collected and employed each of the seven model development approaches described above to evaluate the impact of data sharing restrictions on the performance of ML models.

To isolate the contribution of site size to FL performance, we used the same data as in the primary analysis but randomly allocated all records into groups of the same sizes as the sites in CCASAnet. This created six simulated sites that were independent and identically distributed (IID) samples of the overall CCASAnet cohort. This process effectively eliminated heterogeneity between sites, thereby ensuring that the size of each site (i.e., number of PLWH) was the only meaningful difference between sites that could contribute to performance differences. Site sizes were kept identical to those in the primary analysis to allow for direct comparisons.

To directly assess the impact of between-site heterogeneity, we applied a latent-variable clustering algorithm, Latent Dirichlet Allocation (LDA), to split the data from one large CCASAnet site (Brazil) into three subgroups, a procedure commonly used to study FL under data heterogeneity.^32^ We then sampled from these subgroups to create simulated sites of varying heterogeneity, controlled by a heterogeneity parameter α. A value of α = 0 yields sites that are homogenous with respect to the population, whereas α = 1 yields sites that are as heterogeneous as permitted by LDA. In this situation, the sites are the same as the LDA clusters. We varied α to generate datasets of varying heterogeneity and employed each of the seven ML training algorithms, measuring the performance of each.

### Metrics & Evaluation

To ensure adequate data were available from each site for training and evaluation, we split the data into training, validation, and test sets by thirds, repeating this process over 250 random dataset splits. We evaluated model performance using area under the receiver operating characteristic curve (AUC), sensitivity, specificity, and F1 score, all computed on the held-out test set. Predictive error on the validation set was used to halt model training to prevent over-fitting. The decision threshold to convert probabilities to binary predictions was selected as the threshold that maximized the F1 score on the validation set. For methods that developed one model per site (namely Centralized-FT, FedAvg-FT, FedProx-FT, and Site-Specific), we tuned and applied decision thresholds within each site. By contrast, Centralized, FedAvg, and FedProx relied on a single decision threshold for all sites. Additionally, overall performance measures for FedAvg-FT, FedProx-FT, and Site-Specific methods were computed by pooling the test set predictions from each site and then computing the aggregate AUC, sensitivity, specificity, and F1. We report AUC as our primary performance measure and report the other performance measures in the Supplementary Figures 1-3. We computed averages and standard errors (SE) for each of our performance measures.

## Data Availability

The de-identified data used in this study are available upon the submission and approval of a CCASAnet concept sheet. Additional instructions are available at www.ccasanet.org.

